# Neonatal outcomes following a preconception lifestyle intervention in people at risk of gestational diabetes: secondary findings from the BEFORE THE BEGINNING randomised controlled trial

**DOI:** 10.1101/2025.10.01.25337073

**Authors:** MAJ Sujan, HMS Skarstad, G Rosvold, SL Fougner, T Follestad, SA Nyrnes, KÅ Salvesen, T Moholdt

## Abstract

**Objectives:** Gestational diabetes mellitus (GDM), particularly when combined with overweight or obesity, is associated with adverse neonatal outcomes such as high birth weight and increased adiposity. We determined the effect of a preconception lifestyle intervention initiated before and continued throughout pregnancy on neonatal, birth-related, and body composition outcomes at birth and 6–8 weeks of age in children of participants in the BEFORE THE BEGINNING randomised controlled trial.

**Methods:** People (N = 167) at increased risk of GDM and planning pregnancy were randomly allocated 1:1 to intervention or control. The intervention included time-restricted eating and exercise training. Time-restricted eating involved consuming all energy within ≤ 10 hours/day, ≥ 5 days per week and the amount of exercise was set using a heart rate-based physical activity metric (Personal Activity Intelligence, PAI), with the goal of ≥ 100 weekly PAI-points. The primary outcome in this report was the proportion of infants with birth weight > 4.0 kg.

**Results:** Among 106 live births, 21% (11/53) of infants in the intervention group and 28% (15/53) in the control group had birth weight > 4 kg (*p* =.367). Mean birth weight did not differ significantly between groups (mean difference -159.3 g, 95% confidence interval - 375.7 to 57.2, *p* =.148). No significant between-group differences were found for additional neonatal, birth-related, or early postnatal body composition outcomes.

**Conclusion:** In this secondary analysis, a preconception lifestyle intervention did not reduce the risk of macrosomia or affect neonatal body composition.

**Trial registration:** ClinicalTrials.gov NCT04585581.

## INTRODUCTION

The prevalence of overweight and obesity has risen sharply worldwide. Currently, about 44% of adult women are overweight or obese[1], conditions that increase the risk of pregnancy complications, especially gestational diabetes mellitus (GDM)[2]. GDM, particularly when combined with elevated maternal body mass index (BMI), is associated with adverse perinatal outcomes, including fetal overgrowth, neonatal obesity, and obstetric complications, including preterm birth, shoulder dystocia, neonatal asphyxia, induction of labour, caesarean or instrumental delivery, and postpartum haemorrhage.[3–6] Moreover, offspring of affected pregnancies are predisposed to long-term health consequences, including childhood and adolescent obesity and type-2 diabetes.[7,8]

While exercise-only interventions have been associated with a 39% reduction in odds of macrosomia (birth weight> 4kg)[9], most systematic reviews and meta-analyses conclude that lifestyle interventions in pregnancy have little effect on birth weight or other neonatal outcomes.[9–15] Notably, most lifestyle interventions have been initiated at gestational weeks 16-20, missing the preconception and early gestational window critical to optimise maternal and fetal health.[16] Observational evidence suggests that a healthy maternal lifestyle before pregnancy benefits embryonic development and long-term offspring health.[16] Nevertheless, few randomised controlled trials (RCTs) have investigated the effects of preconception lifestyle interventions on neonatal outcomes[17–20], with mixed results: one RCT demonstrated reduced proportion of neonates born large for gestational age (LGA) in the intervention group[17], whereas others reported no effect[18–20].

The BEFORE THE BEGINNING trial [21] included participants at increased risk of GDM prior to conception. The participants received a combined time-restricted eating and exercise intervention, which was continued throughout pregnancy. While the intervention did not improve maternal glucose tolerance, it reduced weight and fat mass by gestational week 28.[22] Here we report secondary neonatal and birth-related outcomes. We hypothesised that the intervention would reduce the incidence of birth weight > 4 kg and improve additional birth-related outcomes.

## METHODS

### Study design and setting

The BEFORE THE BEGINNING was a single-centre randomised controlled trial conducted at the Norwegian University of Science and Technology (NTNU) in collaboration with St. Olav’s Hospital in Trondheim, Norway. We registered the trial in ClinicalTrials.gov (NCT04585581) on September 25^th^, 2020. All participants signed an informed written consent. A detailed study protocol, including modifications made after trial commencement [21], as well as the primary findings [22] have been published previously. Figure 1 provides an overview of the study design.

**Figure 1:**
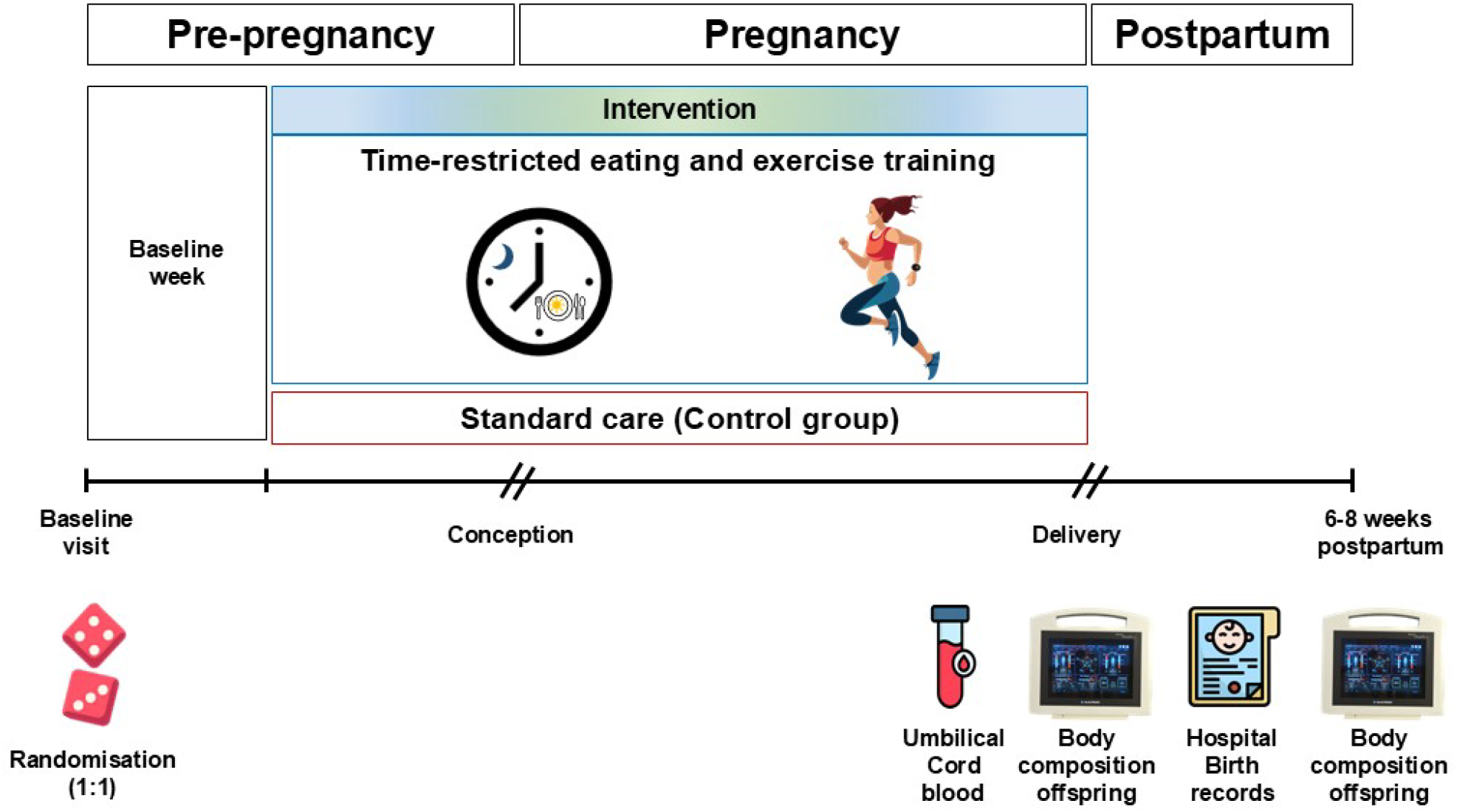
Study design. After baseline assessments, the participants were randomly allocated (1:1) to a lifestyle intervention or a standard care control group. The intervention consisted of time-restricted eating and exercise training, started before, and continued throughout pregnancy. We obtained birth weight and secondary neonatal and birth-related outcomes from hospital records and collected umbilical cord blood samples immediately after delivery. We assessed infant body composition within 72 hours after birth and at 6-8 weeks of age.

### Recruitment and participants

We recruited participants through advertisements on social media, hospital and university websites, local stores, and public spaces. In November 2022, we expanded recruitment by using population data from the Norwegian Tax Administration to distribute electronic invitations. Eligible participants were females aged 18-39 years who planned to conceive within 6 months, understood oral and written Norwegian or English, and met the Norwegian guidelines’ criteria for GDM risk. Our exclusion criteria were: ongoing pregnancy, trying to conceive ≥ 6 cycles, known diabetes (type 1 or 2), shift work involving night shifts > 2 days/week, a history of hyperemesis, known cardiovascular diseases, high-intensity exercise > 2 times/week in the last 3 months, habitual eating window ≤ 12 hours/day, previous bariatric surgery, or any other reason which according to the researchers made the potential participant ineligible.

### Randomisation and blinding

After baseline assessments, we randomly allocated the participants (1:1) to the intervention or a standard care control group, stratified by GDM in a previous pregnancy (yes/no). Details about methods used for random sequence generation, concealment, and block sizes were published previously.[21,22] Neither the participants nor the investigators were blinded to randomisation because of the nature of the intervention. Outcome assessors were not blinded.

### Intervention and adherence

Participants in the intervention group underwent a lifestyle program combining time-restricted eating and exercise training, initiated before conception and continued throughout pregnancy. Time-restricted eating limited daily energy intake to ≤ 10 hours, ending by 19:00, on ≥ 5 days per week. Non-caloric beverages were allowed outside the eating window. We asked the participants to record their eating window in a study handbook every 8^th^ week. Exercise training was guided by Personal Activity Intelligence (PAI), a heart rate-based measure of physical activity.[23] We instructed the participants to maintain ≥ 100 weekly PAI points through high-intensity endurance exercise, adapted for pregnancy. For details on the exercise intervention, see the study protocol.[21] Training was primarily unsupervised, supplemented by supervised sessions at 2 and 8 weeks after baseline and on request, and adherence was monitored via smartwatches (Amazfit GTS/Polar Ignite 2) transmitting data to the research team. Control participants received standard care and were asked to maintain their habitual physical activity and diet.

### Experimental procedures and outcome measures

Here, we report neonatal and birth-related outcomes and infant body composition within 72 hours of birth and at 6-8 weeks. The primary outcome was the frequency of birth weight > 4 kg. From hospital records, we obtained mode of delivery, perineal tear, episiotomy, shoulder dystocia, postpartum haemorrhage, and duration of hospital stay. Neonatal outcomes included birth weight, length, head circumference, APGAR score, and gestational age at birth. The APGAR score is a standardised tool to assess neonatal health immediately after birth, comprising five criteria (skin colour, heart rate, reflexes, muscle tone, and respiration), each scored from 0 to 2, with 10 as maximum. We report the frequency of APGAR score < 7 at 5 minutes, as values 7-10 are considered normal[24]. Preterm birth was defined as delivery before 37 gestational weeks.

Midwives collected umbilical cord blood immediately after birth, before delivery of the placenta. After resting in room temperature for 30 minutes, samples were centrifuged (3100 RPM, 18 °C, 10 minutes), aliquoted, and stored at -80 °C until analysis. Serum insulin C-peptide was measured by electrochemiluminescence immunoassay (Roche Cobas Pro e801), and glucose by photometric assay (Siemens Atellica CH930), both at the St. Olavs’s Hospital laboratories.

We estimated infant body composition using bioelectrical impedance analysis (BIA, BioScan touch i8-nano, Maltron, UK) within 72 hours of birth and at 6-8 weeks of age. Before the measurement, we entered gestational age at birth, age at measurement, sex, ethnicity, length, and body weight into the Bioscan device. Trained staff placed electrodes on the dorsal surface of the right wrist, metacarpal, ankle, and metatarsal while infants lay supine. The device passes a low-amplitude, multi-frequency electrical current through the body and calculates resistance and reactance values to estimate total body water, fat-free mass, fat mass, and muscle mass using age- and sex-specific equations. Trained study personnel performed all measurements to ensure accuracy and repeatability. The measurements were non-invasive, brief, and well tolerated.

### Statistical analysis

We calculated the sample size for the BEFORE THE BEGINNING trial based on the primary outcome measure: 2-hour plasma glucose concentration during a 75 g oral glucose tolerance test (OGTT) in gestational week 28. To detect a between-group difference of 1.0 mmol/L, with a standard deviation (SD) of 1.3, 90% power, and a two-sided significance level of.05, we needed 74 participants. To account for ∼50%, non-conception, 10-20% drop-out, and to increase statistical power for secondary analysis, we initially aimed to enrol 260 participants. Recruitment stopped at 167 participants, deemed sufficient to offset attrition and non-conception.[21] No separate sample size calculation was performed for the secondary outcomes reported here.

We performed intention-to-treat analyses including 106 participants who gave birth. We compared continuous outcomes at birth using two-tailed independent t-tests or Mann-Whitney U-tests and categorical outcomes using χ^2^ or Fisher’s exact tests. We assessed data normality using the Shapiro-Wilk test and QQ plots. Estimated effects are reported as mean differences or risk ratios in the intervention group compared with the control group, with corresponding 95% confidence intervals (CIs) and *p*-values. Rare categorical outcomes are presented descriptively. We evaluated changes in infant body composition from birth to 6-8 weeks using linear mixed models with time, group, and group x time interaction as fixed effects and participant ID as a random effect.

To reduce false positives from multiple secondary outcomes, we set significance at.01. We also performed per-protocol analyses according to the prespecified statistical analysis plan.[21] The statistical analyses were performed using SPSS v29.0.

### Patient and public involvement

We involved users (reproductive-aged women with overweight/obesity) in the planning and implementation of the study. Participant representatives were not involved in the recruitment process. We organised multiple interactive digital meetings and workshops to discuss barriers to participation, motivation, engagement, data collection methods, recruitment strategies, and adherence throughout the study.

## RESULTS

We enrolled 167 participants between October 2^nd^, 2020 and May 12^th^, 2023. Within the specified time frame, 111 participants became pregnant (control, *n = 55, inter*vention, *n* = 56, Figure 1). We excluded data from one participant in the intervention group due to prepregnancy diabetes and one control participant due to a twin pregnancy. Neonatal and birth-related outcomes were missing for three participants. Two participants in the intervention group experienced spontaneous abortion, and one in the control group underwent induced abortion. In total, we included data from 106 live births (control, *n = 53, inter*vention, *n = 53) in th*e intention-to-treat analyses of neonatal and birth-related outcomes (Figure 2). The baseline characteristics were comparable between groups (Table 1).

**Figure 2:**
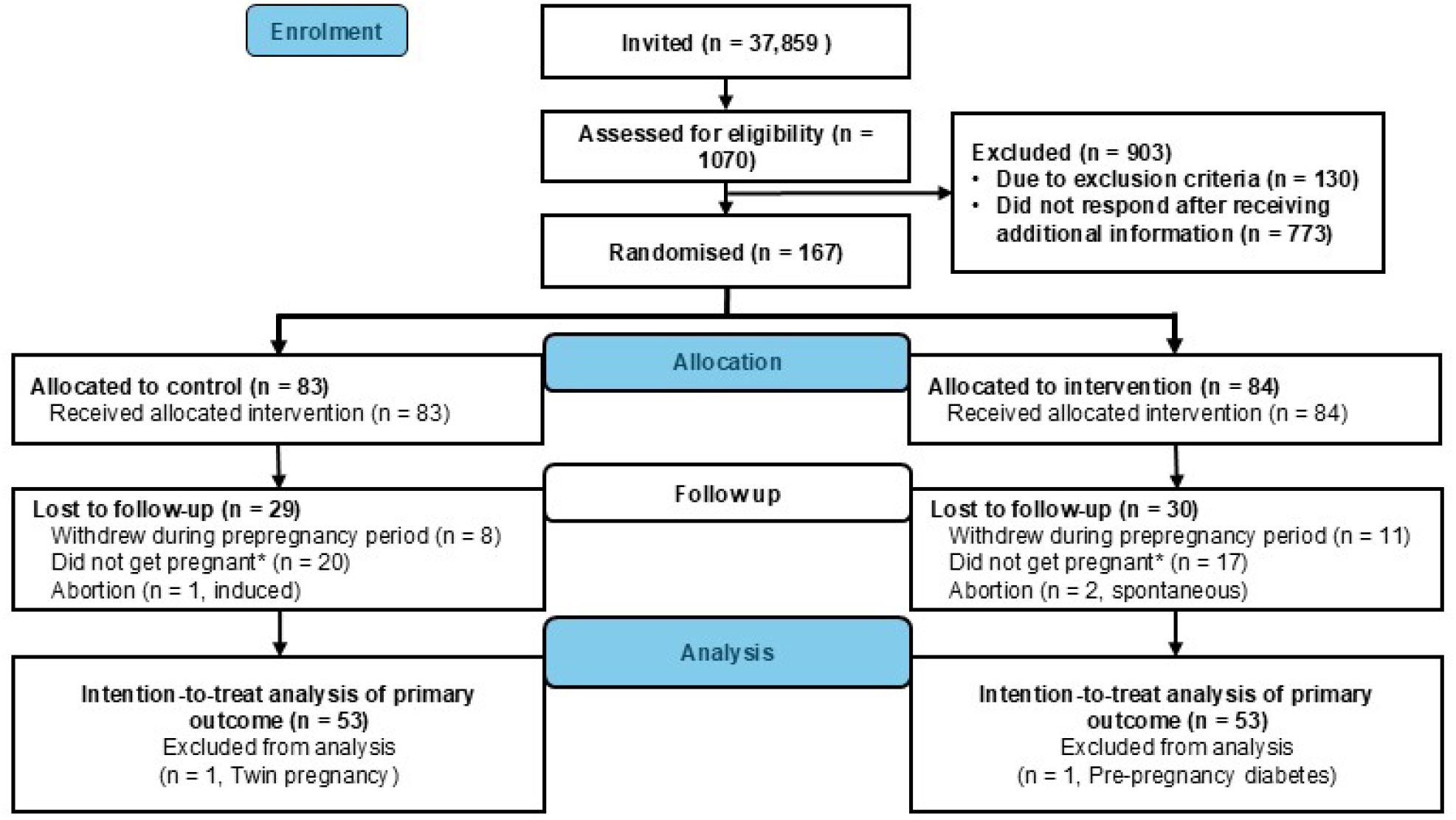
Flowchart of participants (CONSORT Flow diagram).

**Table 1.** Baseline characteristics of participants who delivered a singleton live birth, according to group allocation.

|  | Control (n = 53) | Intervention (n = 53) |
| --- | --- | --- |
| Age, years | 29.5 (3.1) | 29.8 (3.4) |
| Weight, kg | 80.0 (12.8) | 80.8 (15.9) |
| Body mass index, kg/m<sup>2</sup> | 28.5 (4.4) | 29.1 (4.9) |
| Waist circumference, cm | 93.3 (11.2) | 92.5 (12.9) |
| Fat percentage, % | 37.1 (7.1) | 36.3 (7.9) |
| Systolic blood pressure, mmHg | 120 (10) | 119 (8) |
| Diastolic blood pressure, mmHg | 79 (7) | 79 (6) |
| HbA1c, mmol/mol | 34.1 (2.8) | 34.3 (2.8) |
| Fasting glucose, mmol/L | 5.0 (0.4) | 5.0 (0.4) |
| Total cholesterol, mmol/L | 4.5 (0.8) | 4.6 (0.8) |
| HDL cholesterol, mmol/L | 1.4 (0.3) | 1.4 (0.3) |
| LDL cholesterol, mmol/L | 3.0 (0.8) | 3.0 (0.8) |
| Triglycerides, mmol/L | 1.0 (0.5) | 1.0 (0.6) |
| **Parity, total number of births, n (%)** |  |  |
| 0 | 27 (51) | 27 (51) |
| 1 | 20 (38) | 23 (43) |
| 2 | 6 (11) | 3 (6) |
| **Education level, n (%)** |  |  |
| Completed compulsory schooling and upper secondary school | 5 (9) | 10 (19) |
| Completed university education, less than 4 years | 18 (34) | 14 (26) |
| Completed university education, 4 years or more | 30 (57) | 29 (55) |
| **Ethnic origin, n (%)** |  |  |
| Europe | 47 (89) | 47 (89) |
| Africa and Middle East | 2 (4) | 0 (0) |
| Asia | 3 (6) | 4 (8) |
| North America | 0 (0) | 1 (2) |
| Latin America | 1 (2) | 1 (2) |
| **Reason for inclusion\*, n (%)** |  |  |
| Body mass index ≥ 25 kg/m<sup>2</sup> | 45 (85) | 44 (83) |
| GDM in a previous pregnancy | 1 (2) | 1 (2) |
| Family history of diabetes, *n* | 14 (26) | 15 (28) |
| Previous newborn > 4.5 kg, *n* | 0 (0) | 1 (2) |

Data are presented as means of observed values with standard deviation (SD) or number (n) and percentage (%) of participants. Missing: number of participants with missing data in each group is 0 to 4 for all variables. HbA1c = Glycated haemoglobin, HDL = High-density lipoprotein cholesterol, LDL = Low-density lipoprotein cholesterol, GDM = Gestational diabetes mellitus. \*Some fulfilled more than one reason.

### Neonatal outcomes

In the intervention group, 21% (11/53) of newborns had a birth weight > 4 kg, compared with 28% (15/53) in the control group (*p* =.367, Table 2). Birth weight did not differ significantly between groups (mean difference -159.3 g, 95% CI -375.7 to 57.2, *p* =.148). We found no statistically significant between-group differences in other neonatal outcomes or in body composition outcomes assessed within 72 hours of delivery (Table 2). One newborn in the control group had an APGAR score < 7 at 5 minutes after delivery, whereas none in the intervention group did (Table 2). We estimated infant body composition at a mean of 33 hours (SD 1.1) after delivery and again at 7 weeks (SD 1.4). From birth to 6-8 weeks, there were no statistically significant between-group differences in changes in infant body weight, fat mass, fat-free mass, percentage of fat mass and fat-free mass, muscle mass, or hydration (Figure 3 and Supplementary table 1).

**Table 2.** Neonatal outcomes at birth.

| Outcome | Control (*n* = 53) | Intervention (*n* = 53) | Estimate | 95% CI | *p* |
| --- | --- | --- | --- | --- | --- |
| Weight, g | 3702.5 (517.8) | 3543.3 (602.7) | -159.3 | -375.7 to 57.2 | .148<sup>a</sup> |
| Length, cm | 50 (49, 51) | 50 (49, 52) | - | - | .930<sup>b</sup> |
| Head circumference, cm | 36.0 (35, 37) | 36 (35, 37) | - | - | .302<sup>a</sup> |
| Birth weight > 4 kg, *n* (%) | 15 (28) | 11 (21) | 0.7 | 0.4 to 1.4 | .367<sup>c</sup> |
| APGAR < 7 at 5 minutes, *n* (%) | 1 (2) | 0 (0) | - | - | - |
| Preterm birth, *n* (%) | 1 (2) | 2 (4) | - | - | - |
| Gestational age at birth, weeks + days | 40+3 (39+2, 41+1) | 40+0 (39+0, 41+0) | - | - | .319<sup>b</sup> |
| Serum glucose, mmol/L* | 5.6 (1.3) | 5.6 (1.3) | 0.0 | -0.6 to 0.6 | .997<sup>a</sup> |
| Serum insulin C-peptide, nmol/L* | 0.5 (0.5, 0.7) | 0.5 (0.5, 0.6) | - | - | .367<sup>b</sup> |
| Fat mass, kg** | 0.2 (0.1, 0.3) | 0.2 (0.1, 0.3) | - | - | .690<sup>b</sup> |
| Fat mass, %** | 4.8 (2.9, 7.2) | 5.5 (3.0, 7.6) | - | - | .622<sup>b</sup> |
| Fat-free mass, kg** | 3.5 (1.5) | 3.3 (1.5) | -0.1 | -0.3 to 0.1 | .218<sup>a</sup> |
| Fat-free mass, %** | 95.2 (92.9, 97.0) | 94.7 (92.5, 97.0) | - | - | .605<sup>b</sup> |
| Muscle mass, kg** | 1.1 (0.2) | 1.1 (0.2) | -0.1 | -0.1 to 0.0 | .196<sup>a</sup> |
| Hydration (TBW), %** | 79.6 (76.3, 81.8) | 78.6 (75.6, 81.2) | - | - | .449<sup>b</sup> |

Data are observed mean and standard deviation (SD) or median and 25- and 75-percentiles (quartiles) or frequency and percentage. Results are presented as mean differences or relative risks (RR) with corresponding 95% confidence intervals (CI) and *p*-values. TBW = Total body water.
<sup>a</sup>Independent samples t-test
<sup>b</sup>Mann-Whitney U-test
<sup>c</sup>Chi-Square test
\*Control, *n* = 31, Intervention, *n* = 37
\*\*Control, *n* = 49, Intervention, *n* = 45

**Figure 3.**
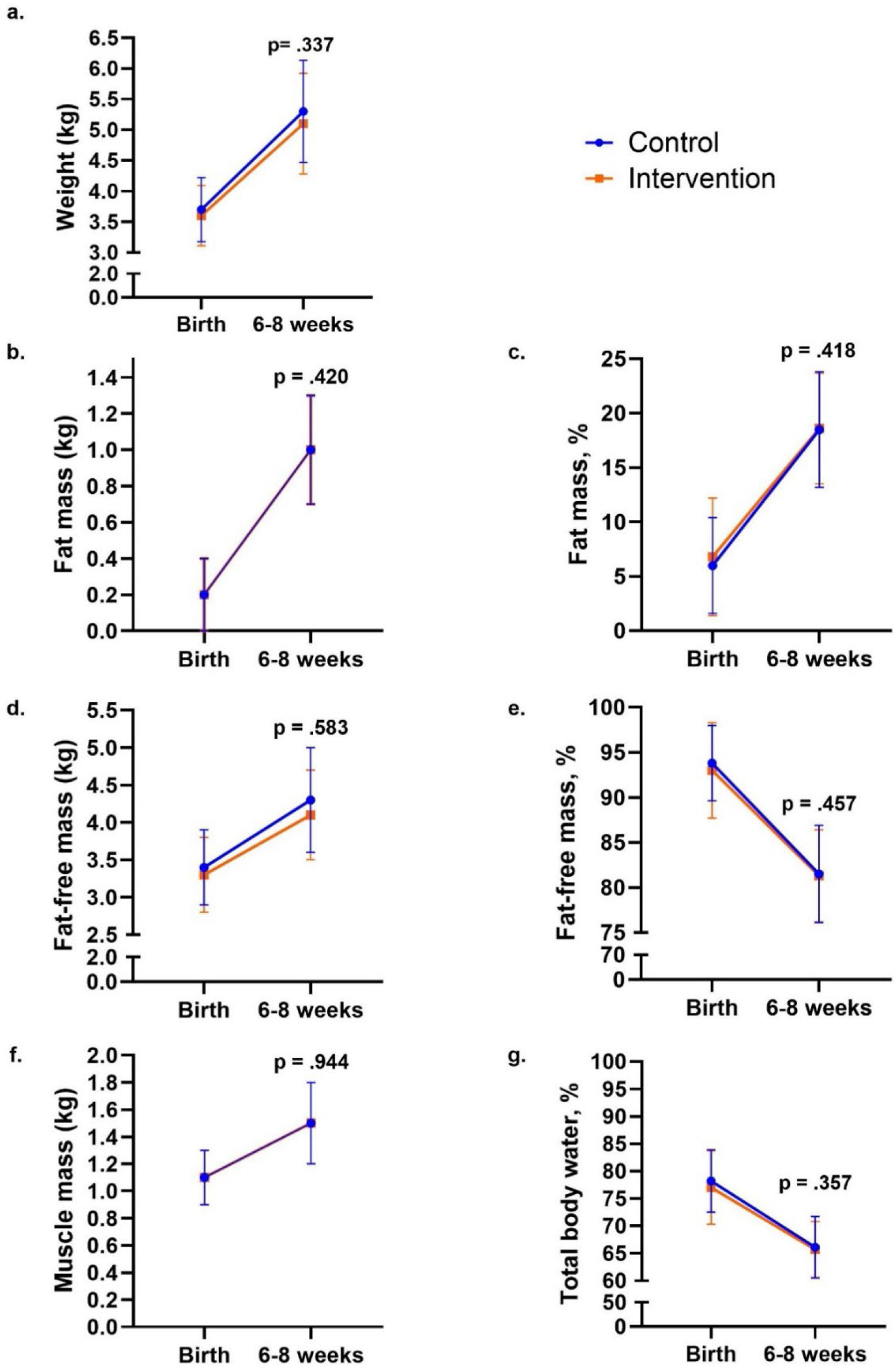
Body composition outcomes of infants at birth and 6-8 weeks of age according to group. a. Body weight, b. fat mass, c. percentage of fat, d. fat-free mass, d. percentage of fat-free mass, e. muscle mass, f. hydration (total body water percentage). Data are observed means and error bars show mean ± 1 standard deviation (SD). The lines represent changes in each group from birth to 6-8 weeks of age. The *p-*values were calculated for between-group differences using linear mixed model.

### Birth-related outcomes

There were no statistically significant differences between groups in mode of delivery, perineal tear, episiotomy, postpartum haemorrhage, or length of the hospital stay (Table 3). One case of shoulder dystocia occurred in the control group, and none in the intervention group (Table 3). Nine of 53 participants (17%) in the intervention group and four of 53 (8%) in the control group were delivered by caesarean section (relative risk 2.3, 95% CI 0.7 to 6.9, *p* =.139). The median length of hospital stay was three days in both groups.

**Table 3.** Birth-related outcomes.

| Outcome | Control<br>(*n* = 53) | Intervention<br>(*n* = 53) | Relative risk | 95% CI | *p* |
| --- | --- | --- | --- | --- | --- |
| Normal vaginal delivery, *n* (%) | 47 (89) | 42 (79) | 0.9 | 0.8 to 1.1 | .186<sup>b</sup> |
| Induced labour, *n* (%) | 10 (19) | 11 (21) | 1.1 | 0.5 to 2.4 | .807<sup>b</sup> |
| Instrumental delivery*, *n* (%) | 2 (4) | 2 (4) | - | - | - |
| Caesarean section, *n* (%) | 4 (8) | 9 (17) | 2.3 | 0.7 to 6.9 | .139<sup>b</sup> |
| Perineal tear*, grade 3-4, *n* (%) | 1 (2) | 1 (2) | - | - | - |
| Episiotomy*, *n* (%) | 5 (10) | 2 (5) | 0.4 | 0.1 to 2.2 | .440<sup>a</sup> |
| Shoulder dystocia, *n* (%) | 1 (2) | 0 (0) | - | - | - |
| Postpartum haemorrhage, *n* (%) | 8 (15) | 6 (11) | 0.8 | 0.3 to 2.0 | .566<sup>a</sup> |
| Length of hospital stay, days | 3 (2, 4) | 3 (2, 4) | - | - | .247<sup>c</sup> |

Data are presented as frequency and percentage or median and quartiles. Results are presented as relative risks (RR), with corresponding 95% confidence intervals (CI) and *p*-values.
<sup>a</sup>Fisher's exact test
<sup>b</sup>Chi-square test
<sup>c</sup>Mann-Whitney U-test
\*Only for vaginal deliveries (control, *n* = 48, intervention, *n* = 44)

### Per protocol analysis

For the per-protocol analysis, we included intervention participants who achieved ≥ 75 weekly PAI-points, adhered to ≤ 10-hour time-restricted eating on at least two of four registered days in each preconception registration, and delivered a live infant. Twenty-three of 53 intervention participants (43%) met these criteria. In total, we included 75 participants (control, n = 52, intervention, n = 23) in the per-protocol analyses. The per-protocol results were consistent with the intention-to-treat findings (Supplementary tables 2-4).

## DISCUSSION

### Main findings and comparison with previous studies

In the BEFORE THE BEGINNING trial, a preconception intervention combining time-restricted eating and exercise training did not significantly affect birth weight, infant body composition, or other birth-related outcomes. Our findings align with previous preconception lifestyle intervention studies in populations at high risk of GDM, reporting no significant impact on neonatal outcomes.[18–20] We observed a small, non-significant reduction in mean birth weight (159 g) in the intervention group compared with the control group, consistent with findings from two prior studies of combined preconception dietary and exercise interventions.[19,20] Maternal prepregnancy BMI and weight gain during pregnancy influence birth weight.[25] In our study, more than 80% of the participants were enrolled due to elevated BMI (≥ 25 kg/m^2^). Although weight loss before or during pregnancy was not specifically targeted, participants in the intervention group gained significantly less body weight and fat mass in late pregnancy.[22] However, these changes may not have been sufficient to improve maternal glycaemic control or other cardiometabolic outcomes influencing neonatal outcomes.

Unlike our trial, previous preconception trials have emphasised weight loss before pregnancy and gestational weight maintenance.[17–20] Notably, a 12-week very-low-energy diet intervention reduced the prevalence of LGA infants from 24% in the control group to 4% in the intervention group among participants with a BMI between 30 and 55 kg/m^2^. This intervention, consisting of a preconception very low-energy diet (800 kcal/day) for 12 weeks, followed by an energy-balanced diet combined with physical activity (> 10,000 steps/day), induced significant weight loss before conception.[17] The resulting improvement in maternal metabolic status may have optimised the intrauterine environment, thereby decreasing the risk of LGA births in the intervention group.[17]

Lifestyle interventions initiated during pregnancy have produced inconsistent effects on birth weight outcomes. A meta-analysis showed that exercise-only interventions were more effective than combined exercise and co-interventions, reducing the odds of birth weight > 4kg by 39%.[9] Most studies included in that meta-analysis relied on unsupervised exercise or counselling-based approaches, which were associated with lower adherence. The participants in our trial adhered well to the intervention during the preconception period, however, adherence declined as pregnancy progressed.[22]

Our study, consistent with two previous preconception trials and several pregnancy lifestyle intervention studies, did not demonstrate statistically significant effects of the intervention on adverse neonatal or birth-related outcomes.[9,10,18,20,26–28] In contrast, the very low-energy diet intervention study reported a significantly lower risk of a composite of obesity-related adverse pregnancy outcomes in the intervention group compared with the control group.[17] The overall incidence of adverse neonatal and birth-related outcomes in our study was low, which limits the statistical power to detect meaningful between-group differences. Complications such as shoulder dystocia, low APGAR scores, preterm birth, episiotomy, caesarean section, and postpartum haemorrhage occurred infrequently in both groups. This lower incidence may reflect differences in clinical practice, participant characteristics, or the relatively healthy baseline status of our cohort despite an elevated GDM risk.

Cord blood glucose and insulin C-peptide concentrations reflect maternal metabolic status and glycaemic and insulin homeostasis during pregnancy.[29] Consistent with the findings from the LIMIT[30] and UPBEAT trials[31], we found no statistically significant between-group differences in cord blood glucose or insulin C-peptide concentrations.

There were no significant between-group differences in infant body composition outcomes measured within 72 hours of birth, consistent with the conclusions of two meta-analyses of lifestyle interventions during pregnancy.[9,25] Of note, the estimated neonatal mean fat percentage in our cohort was lower (6.4%) than that reported in several other lifestyle interventions in women with overweight/obesity (10-14%)[32–36], which could potentially be attributed to the use of BIA. BIA is safe, cost-effective, rapid, and practical for longitudinal measurements, but tends to underestimate fat mass and overestimate fat-free mass compared with air-displacement plethysmography and dual-energy X-ray absorptiometry in paediatric populations.[37] Despite this limitation, we aimed to obtain longitudinal body composition measurements and compare changes between groups.

Infant body composition outcomes did not change significantly between groups at 6-8 weeks of age. Our pooled data indicated a three-fold increase in the percentage of body fat from birth, accounting for □ 47% of total weight gain. Early infant feeding practices influence growth trajectory and body composition; exclusively breastfed infants show a greater increase in body fat percentage than formula-fed infants.[38–41] In our study, breastfeeding practices were similar between the groups and a high proportion of infants (35/42, 83%) were exclusively breastfed, which may have contributed to the observed increase in fat percentage. In contrast, in an Australian cohort of infants born to mothers with healthy BMI and no GDM, fat percentage doubled after 6 weeks, contributing to 40% of total weight gain[38]. They observed no difference between breastfed and other infants in fat-mass increase as a proportion of weight gain at 6 weeks and 3 months, but exclusively breastfed infants had a greater increase at 4.5 months. Infant body composition changes rapidly, and accelerated fat-mass gain and rapid growth during this period are associated with increased risk of childhood obesity,[42] suggesting that early growth patterns influence long-term health. Follow-up of these children may clarify the impact of early body composition changes on future cardiometabolic health.

## Limitations

A major limitation of the study was declining adherence during pregnancy despite good adherence during the preconception period.[22] The analyses of neonatal and birth-related outcomes were secondary, and the incidence of some outcomes was very low. Furthermore, we calculated the sample size based on the primary outcome of the BEFORE THE BEGINNING trial, not the secondary outcomes reported here, which increases the risk of Type II errors and may have obscured true effects. We estimated body composition at two time points in early infancy, providing a longitudinal perspective rather than a single cross-sectional measurement. However, the use of BIA is a limitation due to its lower precision than gold-standard methods for assessing body composition.[43]

## Conclusion and implications for clinicians and policymakers

The BEFORE THE BEGINNING trial preconception intervention combining time-restricted eating and exercise training had no statistically significant effect on neonatal outcomes, including birth weight, body composition at birth or 6–8 weeks of age, or other birth-related outcomes. Although the intervention led to beneficial changes in maternal body composition during pregnancy, these changes did not translate into measurable improvements in neonatal or birth-related outcomes. Given the rising prevalence of obesity and diabetes, continued investigation into preconception lifestyle interventions remains a public health priority. Future studies should aim to include more diverse populations, stratify participants by BMI and metabolic risk, and ensure sufficient statistical power to analyse neonatal outcomes. Longitudinal follow-up beyond infancy will be critical to determine whether early lifestyle interventions confer sustained benefits for maternal and offspring

**What is already known on this topic**

- While exercise only interventions in mothers during pregnancy are associated with lower odds of having a newborn with macrosomia, combined lifestyle interventions in pregnancy have minimal impact on neonatal outcomes

**What this study adds**

- In this randomised controlled trial involving mothers at increased risk of gestational diabetes mellitus, a combined preconception lifestyle intervention had no effect on the proportion of babies with birth weight > 4kg and other neonatal, birth-related or body composition outcomes
- Maternal body composition changes may not have been substantial enough to influence neonatal outcomes

**How this study might affect research, practice or policy**

- Prioritising preconception lifestyle interventions with adequate power for secondary analysis and exploring strategies to improve adherence is essential to uncover long-term benefits for maternal and child health health.

## Ethical approval

The Regional Committees for Medical and Health Research Ethics in Norway approved the study (REK 143756).

## Data availability statement

All individual deidentified participant data and statistical codes are available on the Zenodo data repository: 10.5281/zenodo.17130011. For problems accessing the data, contact the corresponding author.

## Author contributions

TM, KÅS, TF, SAN, and SLF conceived and contributed to the study and analysis plan. GR, MAJS, and HMSS collected the data. MAJS and TM analysed the data, and all authors contributed to the interpretation of the data. MAJS drafted the manuscript. All authors reviewed the manuscript, provided feedback and approved the final manuscript.

## Funding and Acknowledgements

The trial is funded by the Novo Nordisk Foundation (NNF19SA058975), The Liaison Committee for education, research, and innovation in Central Norway, and The Joint Research Committee between St. Olavs Hospital and the Faculty of Medicine and Health Sciences, NTNU (FFU). The funding bodies had no role in the design, data collection, analysis, or interpretation of results. The authors are thankful to all the participants for their valuable contributions and the midwives at the Department of Women’s Health, St. Olav’s Hospital, for their valuable assistance in data collection.

